# Taking Control of Your Functional Cognitive Symptoms Workbook: A Novel Intervention

**DOI:** 10.1101/2024.10.15.24315532

**Authors:** E. Cotton, K. L. Mordecai, L. McWhirter, V. Cabreira, R. Van Patten, N.D. Silverberg, A.J. Kaat, W.C. LaFrance

## Abstract

**Background:** Functional cognitive disorder (FCD) is a subtype of functional neurological disorder (FND). FCD can present with various cognitive symptoms, precipitants, and comorbidities (other FNDs, concussion, fatigue, fibromyalgia, etc.). However, there are no widely available existing behavioral health interventions for FCD. The authors’ aim was to develop a therapist-guided treatment for FCD for widespread use among civilians and Veterans.

**Methods:** We adapted a well-known, evidence-based treatment for functional seizures (an adjacent condition often with cognitive symptoms), to fit the hypothesized mechanisms of FCD. The process is presented in GUIDED format (GUIDance for the rEporting of intervention Development). Key processes included determining broad intervention framework, detailed FCD specific content based on expert consensus, evidence, theory, target population centered approaches, specialty subgroup consideration and target population stakeholder feedback for Veteran and civilian populations.

**Findings:** We have created a comprehensive 14-chapter, manualized, therapist-guided neurobehavioral therapy protocol to target FCD symptoms independent of etiology – the *Taking Control of Your Functional Cognitive Symptoms Workbook*. Initial feasibility, tolerability, and utility were completed with 2 target population stakeholders with FCD (one civilian, one Veteran; both PGI-C rating = 1 “Very Much Improved”). The Template for Intervention Description and Replication (TIDieR) checklist is provided as a supplemental table.

**Interpretation:** This new promising multi-modality behavioral health intervention can be considered Stage 1 (i.e. intervention generation, refinement, modification, adaptation, and pilot testing). Further pilot testing is being conducted and will need to be followed by traditional efficacy testing (Stage 2).

**Key Messages:**

- FCD is an increasingly recognized subtype of FND. Despite existing models of FCD mechanisms, there are no widely available dedicated FCD-symptom focused behavioral health interventions.
- We have created the *Taking Control of Your Functional Cognitive Symptoms: Workbook*, a 14-chapter manualized therapist-guided neurobehavioral therapy protocol to target FCD symptoms regardless of cause / associated conditions, which shows early feasibility and promising clinical utility.

## BACKGROUND

### FCD Diagnostic Criteria, Prevalence, Presumed Mechanisms & Treatment Gaps

Functional Cognitive Disorder (FCD) is a common subtype of Functional Neurologic Disorder (FND) in which the most prominent neurological symptoms are cognitive, and are believed to be associated with alterations in the functioning of neural networks rather than identifiable structural lesions^1-3^. Specific diagnostic criteria for FCD have been proposed by Ball et al, 2020^4^ and include: 1) *symptoms* of impaired cognitive function, 2) clinical evidence of internal inconsistency with observed or measured function, or between different situations, 3) symptoms or impairment are not better explained by another medical disorder, and 4) causing substantial distress or impairment in social, occupational, or other important areas of function.

FCD is an umbrella term including patients in which the main focus and cause of disability is distressing cognitive symptoms and preoccupation about cognition, but can also occur within the context of other comorbidities^5-13^. When combined with other comorbidities, such as FND-functional movement or FND-functional seizure subtypes, to meet full criteria for FCD, the cognitive concerns / symptoms alone must cause substantial distress or impairment, beyond the impact of other comorbidities^4^. FCD should be considered within the existing well known heterogeneous and overlapping nature of FND subtypes, precipitating factors, and comorbidities, including viral illness^10^, mild head injury^11,12^, chronic pain / fatigue^8^, mood / psychological factors^14^, among others. Therefore, tolerance for complexity must be built into conceptualization and treatment approaches for FCD^15-17^.

A “classic” case example may be a person with primary / predominant FCD, with rare FND-functional movement or seizure episodes, overlapping anxiety / depression, fatigue, multiple life stresses, and / or remote history of trauma or adverse childhood experiences. Distinctively, this person presents in primary distress over their newly declined and concerning thinking abilities (brain fog, memory concerns, “unable to think anymore”, etc.) to an extent impacting their activities of daily living (ADLs; no longer working, completing their home tasks, socializing, driving, etc.). The relatively infrequent and short duration movement or functional seizure episodes are unpleasant, but relatively tolerable, and not the reason they are primarily distressed and debilitated. They will often acknowledge pre-existing or ongoing comorbidities (anxiety, depression, life stresses, their history factors etc.), but cite these factors never (or rarely) previously disrupted ADLs. It is the “cognitive changes” that are new, concerning, and debilitating.

### Existing Hypothesized FCD Mechanisms

Like other FNDs, approaching FCD conceptualization via a biopsychosocial framework, and consideration of predisposing, precipitating, and perpetuating factors is prudent (See Table 1). It is reasonably hypothesized FCD shares similar *predisposing* and *precipitating* factors to other FND manifestations, which also appears supported by existing FND (FMD, FS)^3,5,14^ and FCD specific literature^1^. While the manifested primary symptom ‘cognitive concerns’ is specific to FCD, the hypothesized *perpetuating* factors also appear consistent with existing FND (FMD, FS) subtypes^18-27^. This suggests possible similar treatment targets for predisposing factors (alexithymia, comorbidities, life stressors), and perpetuating factors (fear, symptom catastrophizing, avoidance). In addition, there is opportunity for symptom specific helpful management strategies, as those with FND commonly employ good intentioned but ultimately unhelpful management strategies prior to treatment.

**Table 1.** Hypothesized FCD Mechanisms.

| Table 1. Hypothesized FCD Mechanisms |  |  |  |
| --- | --- | --- | --- |
| Predisposing Factors | Precipitants | Subjective Cognitive Concerns | Perpetuating Factors |
| • Biological Predisposition<br>• Medical & Psychiatric Comorbidities<br>• Alexithymia<br>• Early Life Experiences<br>• Traumatic Events<br>• Life Stressors | • Viral Illness<br>• Head Injury<br>• Medical Procedure<br>• Medication Change<br>• Information About Risk<br>• Evaluative Circumstance<br>• Normal Aging<br>• Allostatic Load | Examples:<br>• Brain Fog<br>• Memory Failures<br>• Word Finding<br>• Going Blank | • Hyper Arousal<br>• Dissociation<br>• Catastrophic Interpretations<br>• Memory Perfectionism<br>• Hypervigilance To Cognitive Performance<br>• Task Avoidance<br>• Rumination<br>• Excess Compensatory Strategies |
|  |  | Features:<br>• Variable<br>• Internally Inconsistent<br>• Discordant Objective Findings |  |

### Existing Treatment Gaps & Current Study

Table 2 summarizes some of the key existing FND focused behavioral health treatment options. Of note, there are no currently available treatments for FCD that are broadly applicable to FCD (independent of precipitating factor), widely available, or evidence based. However, the *Taking Control of Your Seizures*: *Workbook*^18-20^ has demonstrated early utility of improving subjective cognitive concerns and objective cognition (via the MoCA) in a FS +mTBI sample^21-22^.

**Table 2.** Existing FND Focused Behavioral Health Interventions Versus the *Taking Control of Your Functional Cognitive Symptoms: Workbook*.

|  | Specific to FCD | FCD Regardless of Precipitating Factor or Associated Features | Evidenced Based | Multi-Modal Psychotherapy | Widely Available | Therapist Guided Intervention |
| --- | --- | --- | --- | --- | --- | --- |
| Mindfulness Based Therapy for Psychogenic Non-Epileptic Seizures<sup>24-25</sup> | X | X | ✓ | X | X | ✓ |
| CODES: a multicentre randomised controlled trial (ISRCTN05681227; Completed)<sup>30</sup> | X | X | X* | X | X | ✓ |
| *Overcoming Functional Neurologic Symptoms: A 5 Areas Approach* (Guided Self Help)<sup>29</sup> | X | X | ✓ | X | ✓ | X |
| *Taking Control of Your Seizures Workbook*<sup>31</sup> | X | X | ✓ | ✓ | ✓ | ✓ |
| Feasibility of Cognitive Behavioural Therapy for Functional Memory Symptoms After Concussion (NCT05581810; Completed)<sup>29</sup> | ✓ | X | ✓ * | X | (✓) | ✓ |
| *Taking Control of Your Functional Cognitive Symptoms Workbook* | ✓ | ✓ | -- | ✓ | (✓) | ✓ |

X\*: The CODES (COgnitive behavioural therapy vs standardized medical care for adults with Dissociative non-Epileptic Seizures) trial was not shown to significantly reduce functional seizure frequency, but this was reportedly due to a superior control group intervention; improvements in secondary factors such as mood were significant
✓ \*: This intervention demonstrated significant improvement in cognitive and associated symptoms following concussion, but this improvement was equivalent to the cognitive rehabilitation controls
(✓): Intended for widespread distribution

Other existing treatment options are management of comorbidities, and use of existing (non-FCD targeted) interventions, such as those for anxiety, or objective cognitive impairment. Existing psychotherapy treatment protocols such as the widely used *Unified Protocol for The Transdiagnostic Treatment of Emotional Disorder*^*28*^, are designed to target primary emotional and mood disorders, via teaching behaviors and thoughts that help support and regulate mood. Such tools may be useful for common FCD comorbidities, but do not focus on the cognitive concerns of those with FCD. Further, existing traditional cognitive rehabilitation approaches may seem promising, but they assume clear objective deficit treated by the adoption of compensatory strategies. This poses challenges for those with FCD, as the assumption of objective deficit and over-use of compensatory strategies in some with FCD are hypothesized pathologic mechanisms. However, it may be possible to appropriately modify cognitive rehabilitation for FCD, for some potential benefit, in those with FCD; but this option does not yet exist in any widely available format^29^.

In the current manuscript, we describe the creation of a comprehensive 14-chapter manualized therapist-guided neurobehavioral therapy (NBT) protocol for patients with FCD, targeting core subjective cognitive concerns, regardless of precipitating factor or associated features: the *Taking Control of Your Functional Cognitive Symptoms: Workbook*.

## METHODS

### Generating a Novel FCD Intervention: Taking Control of Your Functional Cognitive Symptoms: Workbook

The novel FCD intervention was developed overall several years through weekly meetings of a multidisciplinary team composed of experts in neuropsychology, neurology, and psychiatry. Figure 1 graphically represents our process. The intervention development is presented using the consensus based “GUIDED” format (GUIDance for the rEporting of intervention Development), a content checklist for intervention development studies in health research proposed by Duncan et al. 2020^32^. The GUIDED format emphasizes the development of the intervention to allow transparency and learning about intervention development research and practice. In brief, the GUIDED reporting items include intervention context, purpose, target population, exiting intervention components utilized, approach to development (evidence, theory, target population centered, etc.), guiding principles, stakeholder involvement, description of iterative process, subgroup changes, and uncertainties of the intervention development.

**Figure 1.**
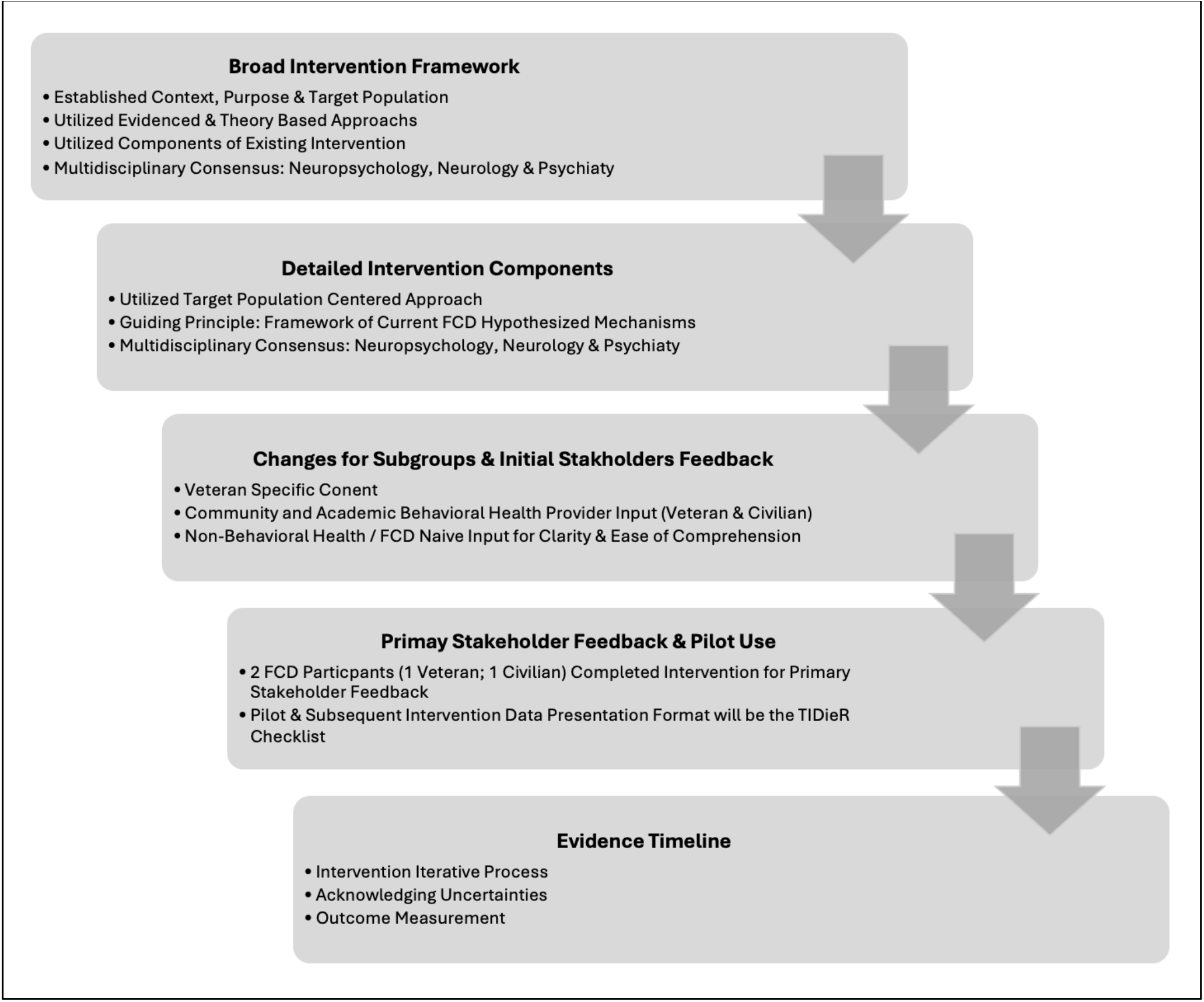
‘GUIDED’ Intervention Development Process.

### Context, Purpose & Target Population of Intervention

The *Taking Control of Your Functional Cognitive Symptoms: Workbook* (*TCY-FCS Workbook*) is intended for use in patients with FCD regardless of precipitating factor or associated features. This intervention is intended to be a consumer publication, in book format, to be easily purchased on-line or in brick-and-mortar bookstores. As such, this intervention is portable, low cost, easy to use, and does not require any specialized equipment. Intended practice settings are wide ranging from large specialized academic medical centers to community and private behavioral health care clinics. Intended clinicians are any licensed mental health care clinician who is tasked with providing behavioral health intervention for a patient with FCD.

The training requirements are intended to mirror that of an existing FND intervention (discussed below), which generally include at a minimum existing knowledge and training in the intervention’s core psychotherapeutic modalities and general awareness of the condition and associated features. Further specialty training can include use of an accompanying ‘therapist guide’ (forthcoming), or in person or remote training of 2 cases by the intervention authors (for supervised training). Additionally, this novel intervention would likely be most easily adopted by those clinicians already acquainted with the approach from the existing “Taking Control Workbook” (see below; Table 3) as the large treatment organization, content flow, session outlines, etc. would be highly familiar, with primarily the FCD specific content being much of the novel substitution. However, lack of familiarity with the existing “Taking Control Workbook” is not expected to preclude successful utilization of this novel intervention for FCD.

**Table 3.** Components of Existing Intervention Utilized *Taking Control of Your Seizures: Workbook* Key Features & Evidence Base Summary.

| **Table 3. Components of Existing Intervention Utilized** |  |
| --- | --- |
| ***Taking Control of Your Seizures: Workbook* Key Features & Evidence Base Summary** |  |
| **Intervention Structure:** | **Theoretical Approach:** |
| • 13 Chapter Workbook<br>• Therapist-Guided / Patient-Led<br>• Intensive & Time Limited<br>• Multi-Modal Intervention<br>• Neurobehavioral Focus | • Biopsychosocial<br>• Biopsychosocialspiritual<br>• Non-Dualist<br>• Integrated Mind-Brain<br>• |
| **Intervention Sequence:** | **Core Therapeutic Modalities:** |
| • Condition Psychoeducation<br>• Overview of Intervention<br>• Introduction to Symptoms and Biopsychosocial Targets<br>• Stepwise Approach to Symptom Pattern Awareness & Management<br>• Use of Weekly Goals to Create Change<br>• Identification of Obstacles to Awareness & Change<br>• Consideration of Symptoms in Holistic Wellness Context<br>• Plan For Continued Progress After Treatment Conclusion | • Psychoeducation<br>• Motivational Interviewing<br>• Cognitive Behavioral Therapy<br>• Acceptance & Commitment Therapy<br>• Dialectical Behavioral Therapy<br>• Agency – Narrative Therapy<br>• Insight Oriented Psychodynamic Therapy |
| **Structure of Intervention Sessions & Approach:** | **Evidence Based For:** |
| • 1 Hour Weekly Appointments<br>• Chapter Read & Homework Completed Prior to Each Appointment<br>• Symptom Log Review & Insight Discussion<br>• GOAT: Goals, Obstacles, Assignments, Tools | • Epileptic Seizure Frequency<sup>33</sup><br>• FS Frequency<sup>18,19</sup><br>• Functional Movement Symptoms<sup>34</sup><br>• Secondary Outcomes of Mood and Quality of Life<sup>18,19</sup><br>• Subjective & Objective Cognition (in FS+mTBI)<sup>23,24</sup> |
| **(For Reference): Workbook Chapters for Seizures / Functional Seizures** |  |
| • Ch 1: Introduction for the Patient: Understanding Seizures<br>• Ch 2: Making the Decision to Begin the Process of Taking Control<br>• Ch 3: Getting Support<br>• Ch 4: Deciding About Your Drug Therapy<br>• Ch 5: Learning to Observe Your Triggers<br>• Ch 6: Channeling Negative Emotions into Productive Outlets<br>• Ch 7: Relaxation Training: Experiencing the Sensation of the Brain Changing Itself | • Ch 8: Identifying Your Pre-Seizure Aura<br>• Ch 9: Dealing With External Life Stresses<br>• Ch 10: Dealing With Internal Issues and Conflicts<br>• Ch 11: Enhancing Personal Wellness<br>• Ch 12: Other Symptoms Associated with Seizures<br>• Ch 13: Taking Control: An Ongoing Process |

The purpose of the intervention is to reduce FCD symptoms and to improve daily functioning and quality of life for individuals with FCD by targeting cognitive, emotional, behavioral and somatic problems common to this presentation phenotype. Importantly, the intervention was specifically developed with language that is specific to FCD, but broad regarding possible precipitating events or factors, or associated features and conditions. Therefore, the *TCY-FCS Workbook* is broadly inclusive of the target population who would meet FCD criteria, and who may have other FNDs, and somatic manifestations, including prolonged post-concussion symptoms, fibromyalgia, persistent symptoms of post viral illness (such as cognitive concerns in the setting of ‘long COVID’) or worried well, among others.

### Evidence & Theory Based Approach for the Broad Intervention Framework & Components of Existing Interventions Utilized

In the first step of intervention development, an evidence- and theory-based approach was used to determine an applicable broad intervention framework. This included overall structure and sequence of the workbook, core therapeutic modalities, and the structure of intervention sessions. The team decided to adapt an existing intervention with solid theoretical approach and robust evidence base, the *Taking Control of Your Seizures: Workbook (TCY-S Workbook)*. This intervention is evidenced based for epilepsy, functional seizures, and functional movement disorder (see below, Table 3).

### Target Population Centered Approach for Detailed Intervention Components & Guiding Principles

Next, after agreement on adopting the broad framework noted above, detailed intervention components (workbook chapters) were adapted using a target population (FCD) centered approach. Co-authors each selected chapters most relevant to their expertise for detailed FCD focused adaptation. See Table 4 for detailed author / chapter listings and contributions. Each author removed / revised any specific seizures / functional seizure content, and added detailed content targeted to an FCD population. Each detailed-content revised chapter was then further reviewed, discussed, and edited in weekly group meetings. A guiding principle during the phase of intervention development was prioritizing close adherence to the existing models of FCD. The FCD hypothesized mechanisms displayed in Figure 1 were used as an anchor point for all chapter detailed edits and larger chapter changes during group discussion of revisions.

**Table 4.**
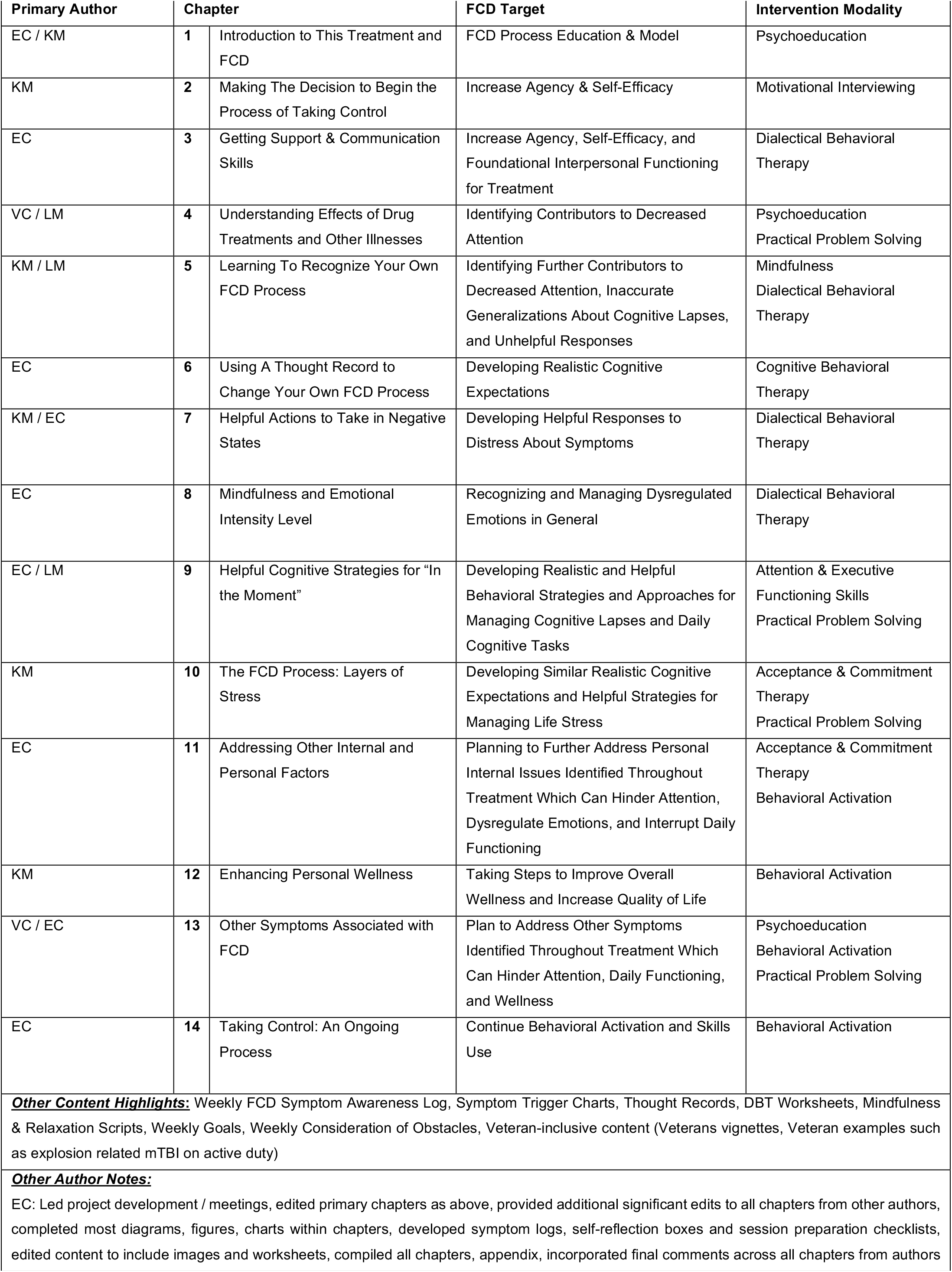
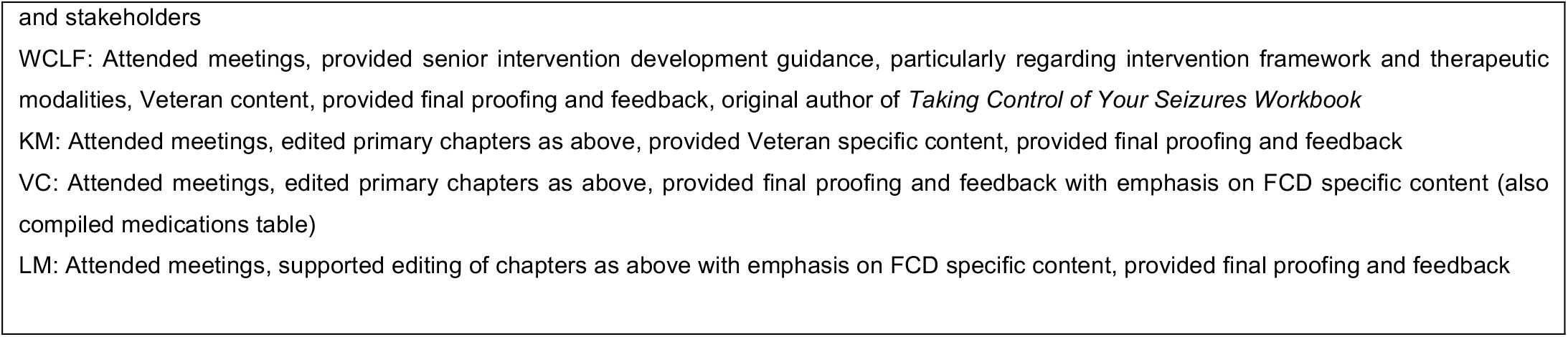
*Taking Control of Your Functional Cognitive Symptoms Workbook* Intervention.

There were three notable large target population centered adaptations for the chapter structure of the workbook. The first being creating a separate chapter for learning and practicing the thought record. In the original *TCY-S Workbook*, the thought record is embedded within another chapter (*TCY-S* Chapter 5). Given the target population (those with FCD), it was decided to create a full separate chapter for slower and more manageable paced approach to learning and practicing this important tool. Second, we expanded the relaxation chapter (*TCY-S* Chapter 7) to include more robust emotional awareness, distress tolerance, and emotional management techniques, along with teaching relaxation strategies. The rationale for this is the hypothesized role of alexithymia traits, hyperarousal and dissociation contributing to FCD symptoms. Thirdly, we heavily adaptated the seizure / functional seizure “pre-seizure aura” chapter (*TCY-S* Chapter 8), to become FCD focused. Specifically, a chapter devoted to helpful strategies, approach, and responses to, cognitively demanding tasks in daily life for people with FCD.

### Changes Required for Subgroups

The original *TYC-S Workbook*, has been widely used with Veterans. As such, we wanted to ensure the intervention adaptation could apply to both civilians and military Veterans with FCD as well. Two of the primary authors (WCL & KM) both practice at U.S. VA Medical Centers, and they led the development of Veterans inclusive content and vignettes, based on their clinical experience with Veterans with FCD. Going forward, with wide-spread use, clinicians already experienced in working with Veterans and Veteran-specific issues would likely maximize this intervention utility for Veterans with FCD. Further, in future intervention trials / use, Veteran specific comorbidities should be characterized and considered (moral injury, head injury, military trauma, etc.) with psychometrically sound and standardized instruments where possible.

## RESULTS

The above process created an intensive, transdiagnostic FCD intervention, the *Taking Control of Your Functional Cognitive Symptoms: Workbook*, with details depicted in Table 4, target population stakeholder feedback presented in Table 5, and TIDieR checklist presented in Supplemental Table 1.

**Table 5.** Target Population Stakeholder Intervention Feedback.

| **Table 5. Target Population Stakeholder Intervention Feedback** |  |
| --- | --- |
| **Broad Intervention Framework** | • Written workbook format was seen as a positive for organization, consistent reference, and post-treatment access to discussed content (empowerment)<br>• Chapter progression and organization was reported as logical, linear, and appropriately scaffolded (foundational at the start, more complex later)<br>• General chapter format was reported as consistent, clear, helpful, and appropriate length |
|  | • Therapist guidance was seen as beneficial (beyond self-help) for optimal implementation of chapter related skills and helpful sense of therapeutic connection<br>• Broad framework elements particularly helpful were self-reflection prompts related to chapter content, use of diagrams to explain content, symptom logs, and thought records |
| **FCD Specific Content** | • A label and description of FCD as a brain-based process was reported as validating and informative<br>• A focus on cognitive symptoms primarily (rather than on mood or physical symptoms) was reported as essential<br>• Description of the FCD process and associated components was reported as a logical and helpful framework to identify aspects of their own FCD process<br>• Blank tables and diagrams for participants own FCD specific elements and process were reported as helpful and organizing<br>• Connecting cognitive issues with other factors (mood/emotions, situations, thoughts, behaviors, physical aspects; a biopsychosocial model) was reported as helpful, sensible, and effective<br>• Emotional awareness and regulation strategies were reported as instrumental as were thought records to challenge unhelpful thinking patterns<br>• Learning normal rates of cognitive lapses, more helpful thoughts in response to cognitive lapses, and drawing attention to cognitive successes were reported as very beneficial |
| **Recommended Changes** | • Minor changes to content clarity were provided for each chapter<br>• Additional skills (grounding strategies for when feeling overwhelmed and a recommended paced workbook approach) were added to Chapter 1 given frequent beneficial use throughout the intervention with target population stakeholders |

### Clinician Stakeholders Feedback

Following full *TCY-FCS Workbook* compiling, clinician-stakeholders provided additional full workbook review and feedback. Specially, feedback was provided from two additional neuropsychology focused behavioral health clinicians who work with a range of emotional and cognitive presentations and concerns --one a civilian in private practice, and one a Veteran clinician in a large academic medical setting. These stakeholders represented the intended behavioral health clinicians who may use this new intervention once it is able to be widely distributed. These clinician-stakeholders contributed edits in the form of detailed workbook review (grammar, errors, content clarity, comprehension level, terminology, etc.) and participated in clinician stakeholder workbook review meetings. Additionally, one non-behavioral health clinician unfamiliar with FCD (a medical student) completed a full workbook review to provide a mental health intervention and process-naïve perspective regarding content clarity, comprehension level, and terminology.

### Target Population Stakeholder Feedback

Two target population stakeholders, one Veteran, one civilian, who both met full criteria for FCD, agreed to clinically complete the entire intervention to incorporate target population stakeholder feedback, and report on preliminary feasibility, tolerability, and utility. Both gave verbal consent for their feedback to be included in this report; also collecting outcomes of clinically based FND interventions is an approved activity by the Northwestern Institutional Review Board. The Veteran patient, with intervention clinician KM at the Baltimore VA, offered target population stakeholder input on a slightly earlier version of the intervention (prior to full incorporation of initial stakeholder input from other clinicians noted above). This Veteran patient-stakeholder was a middle age, graduate level-educated, white, female Veteran, who experienced primary ongoing FCD in the setting of a history of three prior resolved FMD episodes (fatigue, headache, full hemiparesis, numbness), post-traumatic stress disorder, depression, and chronic fatigue syndrome. This target population stakeholder had previously attempted various psychoactive medications, supportive and traditional cognitive behavioral psychotherapy without success. Due to FCD impact, she was on a leave from work, had stopped her hobbies / social activities, and restricted travel. This stakeholder’s intervention was completed via telehealth (stakeholder preference).

The civilian target population stakeholder completed the intervention with clinician EC at Northwestern, using a slightly later version of the intervention (minorly modified version following full clinician stakeholder input from other clinicians noted above). This civilian target population stakeholder was a middle age, college educated, Hispanic, female, with a history of FCD after a Covid-19 infection, with a history of childhood abuse, and anxiety. This patient had previously completed supportive psychotherapy, symptom focused pharmacotherapy, and cognitive rehabilitation with a speech and language pathologist (SLP) without success in symptom reduction. She had discontinued work, driving, and socialization due to FCD symptom interference. This stakeholder’s intervention was completed in person (stakeholder preference).

Both KM and EC recorded detailed intervention notes on each chapter with the target population stakeholders concerning their feedback and suggestions (additional useful skills that were discussed but not explicitly listed in workbook that were then added, sections to clarify / simplify, etc.).

### Target Population Stakeholder Feedback: Preliminary Feasibility, Tolerability, and Utility

The civilian target population stakeholder also completed a modified Treatment Credibility & Expectancy Questionnaire^35^ at the end of Chapter 2 (the chapter that offers a detailed description of the intervention). Further, Patient Global Impression of Change (PGI-C)^36^ was utilized to quantify target population stakeholder reported improvement during the last chapter of the intervention. The PGI-C is a seven-point item scale rating, ranging from 7 (Very Much Worse) to 1 (Very Much Improved).

The civilian target population stakeholder Treatment Credibility & Expectancy Questionnaire responses indicated a 9 regarding perceived intervention credibility (on a scale of 1-9; 9 being the treatment seeming “very logical”), and 9 regarding expectancy of treatment to improve symptoms (on a scale of 1-9; 9 being “very likely” to improve symptoms). Both target population stakeholders took 17 appointments to complete the full 14-chapter *TCY-FCS Workbook* (well within the range as seen in patients with FS, which average 17 sessions^19^). At the time of the last intervention appointment, both target population stakeholders reported a PGI-C rating of 1, “Very Much Improved” (the highest improvement rating). Target population stakeholders described elements believed central to the intervention’s success, reported in Table 5.

## DISCUSSION

### Evidence Timeline, Intervention Iterative Process, and Uncertainties

This novel behavioral health intervention is presently in Stage 1 of the intervention development process (i.e. adaptation and pilot testing). Further pilot testing is being conducted and will need to be followed by traditional efficacy testing (Stage 2). The strengths of this current work include drawing from an evidenced based treatment for another common FND subtype, expert adaptation to target core hypothesized mechanisms of FCD, the inclusion of clinician and target population stakeholder feedback, and demonstration of early feasibility, tolerability, and utility.

However, there are several limitations and present uncertainties in the current work. First, the heterogenous nature of FCD etiology, presentations, and age range suggests a need for consensus, regarding diagnostic criteria, and reliable FCD screening tools or checklists for clinical use with this intervention. Recent literature indicates use of a Delphi study to create an 11-item checklist screening tool for functional memory symptoms^37^. However, it has not yet been validated retrospectively and prospectively for clinical use. At present, minimum standards for clinical and research diagnosis of FCD should include the elements of a standard neuropsychiatric interview, subjective cognitive concern assessment, and objective cognitive measurement to the level necessary for diagnostic confidence. In the interim, for research purposes, blinded review of enrollment decisions (such as 10% of random enrollment sample) could be completed to ensure adequate inter-rater reliability metrics of FCD diagnostic and enrollment decisions.

Also, subjective and objective cognitive clinical outcome assessments need to be evaluated for evidence of sensitivity to meaningful within individual change (change that is meaningful to the patient, not just statistically significant), to be useful in evaluating response to FCD interventions^38-39^. Lastly, FCD intervention trials should include treatment credibility, therapist and patient intervention compliance metrics, consistent with other FND evidenced-based therapies^19,20^.

Given the preliminary nature of this work, completing these steps in future research trials will be an iterative process and essential to moving this intervention into Stage 2 (Efficacy; in a controlled research environment). Future studies may also consider this intervention in a community setting with training material development for real world providers, aimed at optimizing implementability of this intervention in real world settings (Stage 3). This could be followed by fully powered, blinded, randomized clinical trials comparing this intervention to other existing evidenced based traditional manualized behavioral health interventions, cognitive rehabilitation, and pharmacotherapy treatment protocols.

## CONCLUSIONS

We have created a comprehensive, 14-chapter, manualized therapist-guided, patient-led, multi-modality, whole-person, neurobehavioral therapy intervention to target FCD symptoms regardless of precipitating factor or associated features – the *Taking Control of Your Functional Cognitive Symptoms: Workbook*. Broad intervention framework and FCD detailed intervention components were developed based on expert consensus, evidence, theory, and target population centered approaches. Further, changes were incorporated based on specialty patient subgroups (Veterans) and based on feedback from additional clinician stakeholders. Initial feasibility, tolerability, and utility were established with two primary FCD target population stakeholders (one Veteran, one civilian; both with PGI-C rating of 1 “Very Much Improved”). Further pilot data is presently being collected, with plans for research trials to establish appropriate outcome measures and explore efficacy of this novel intervention.

## Data Availability

All data produced in the present study are available upon reasonable request to the authors

## Acknowledgements

We would like to sincerely thank Dr. Sherrie All, Dr. Anna Blanken, and Tali Sorets for their review and feedback of the *Taking Control of Your Functional Symptoms: Workbook* draft. We would also like to thank Dr. Jessica Urgelles for drafting the postconcussive symptoms workbook, Drs. Joel M. Reiter, Donna Andrews, and Charlotte Reiter for their original *Taking Control* Workbook, and permission for adaptation.

