## Supplemental Table 1 for "Taking Control of Your Functional Cognitive Symptoms Workbook: A Novel Intervention"

**Supplemental Information**

| **Supplemental Table 1. Template for Intervention Description and Replication (TIDier) Checklist** | |
| --- | --- |
| **Brief Name** |  |
| 1 | *Taking Control of Your Functional Cognitive Symptoms: Workbook* |
| **Why** |  |
| 2 | Reduce FCD symptoms, improve daily functioning and quality of life for individuals with FCD by targeting cognitive, emotional, behavioral and somatic problems common to this presentation phenotype   - See: Purpose & Target Population of Intervention - See: Evidence & Theory Based Approach for the Broad Intervention Framework & Components of Existing Interventions Utilized |
| **What** |  |
| 3 | Materials: 14 Chapter Workbook   - See: Context of Intervention |
| 4 | Procedures: 14 Chapter Workbook   - See: Table 3 & Table 4 |
| **Who Provided** |  |
| 5 | Licensed Mental Health Care Clinician  Training Requirements; See: Context of Intervention   - Existing knowledge and training in the intervention’s core psychotherapeutic modalities and general awareness of the condition and associated features (See Above: Table 4) - Further specialty training can include use of an accompanying ‘therapist guide’ (forthcoming), or in person or remote training of 2 cases by the intervention authors (for supervised training)   In This Preliminary Use   - Licensed clinical neuropsychologists - Specialty trained in FND / *TCY-S Intervention* |
| **How** |  |
| 6 | Individual Behavioral Health Intervention Sessions  In Person or Telehealth (Clinical Video Telehealth) |
| **Where** |  |
| 7 | With Behavioral Health Providers Tasked with Treating FCD   - See: Context of Intervention   - Intended practice settings are wide ranging from large specialized academic medical centers to community and private behavioral health care clinics   In This Preliminary Use   - Civilian Academic Medical Center - Veterans Affairs Medical Center |
| **When & How Much** |  |
| 8 | 14 Chapter Workbook   - See: Table 3 & 4 - 1 Hour Weekly Sessions   In This Preliminary Use   - Target Population Stakeholders completed 17 total sessions - Consistent with intent for extra weeks on personally relevant chapters / content - This is parsimonious with original *TYC-S Workbook* total sessions |
| **Tailoring** |  |
| 9 | - Adaptations from Original *Taking Control of Your Seizures: Workbook* - See: Figure 1 GUIDED Intervention Development - See: Changes Required for Subgroups (Veterans) - See: Clinician Stakeholder Feedback - See: Target Population Stakeholder Feedback - In This Preliminary Use & Recommended Use Going Forward   - Extra time (3 sessions) were spent on chapters / content most relevant for the person with FCD (common include: thought record, emotion regulation, external stress, internal issues)   - Veteran familiar providers are recommended to administer the intervention to Veterans with FCD |
| **Modifications** |  |
| 10 | Adaptations from Original *Taking Control of Your Seizures: Workbook*   - See: Evidence & Theory Based Approach for the Broad Intervention Framework & Components of Existing Interventions Utilized - See: Changes Required for Subgroups (Veterans) - See: Clinician Stakeholder Feedback - See: Target Population Stakeholder Feedback   In This Preliminary Use & Recommended Use Going Forward   - Additional skills were added to Chapter 1 (grounding strategies for when feeling overwhelmed, and paced approach to workbook reading / homework) which have now been added to the intervention protocol |
| **How Well** |  |
| 11 | Planned: See: Target Population Stakeholder Feedback |
| 12 | Actual: Intervention Delivered as Planned to Population Stakeholders   - Full 14 Chapter Workbook Complete via 17 Weekly Sessions with Population Stakeholders   Future Pilot / Efficacy Studies Should Include:   - Blinded Outcome Assessor for Subjective & Objective Cognitive Outcome Measures - Blinded Enrollment Inter-Rater Reliability for FCD Diagnosis and Enrollment Decisions (such as random %10 of enrollment sample independently reviewed) - Intervention / Therapist Competence & Adherence Checks (such as 2 session random sampling per participant reviewed by trained staff scored for session content adherence) |
